# Cognitive trends in an urban Indian elderly community: Glycated hemoglobin and geriatric depression play a bigger role than age

**DOI:** 10.1101/2023.02.24.23286286

**Authors:** Bhaktee Dongaonkar, Arman Deep Singh, Swathi B Hurakadli, Ashwini Godbole

## Abstract

**Objectives:** To explore the cognitive profile in urban Indian older adults and observe the prevalence of cognitive impairment associated with age, glycated hemoglobin (HbA1c) levels, vitamin B12, and other psychosocial factors

**Methods:** Urban community dwelling older adults (55-85years, n=123) underwent a detailed demographic and cognitive assessment comprising of tests from different cognitive domains – memory, executive function, visuospatial abilities, and verbal fluency. Serum samples were collected from a subset of participants (n=60) to determine HbA1c and vitamin B12 levels.

**Results:** Performance in all cognitive domains declined with age. The decline became prominent around age 70. HbA1c correlated inversely with processing speed and executive function. Vitamin B12 did not correlate with performance on any cognitive test. Geriatric depression correlated inversely with visuospatial abilities. Surprisingly, stepwise multiple regression revealed that HbA1c and geriatric depression contributed to 28% variance on Montreal Cognitive Assessment whereas participant age did not contribute significantly. Mild Cognitive Impairment (MCI) was observed in 17% of participants. Participants classified as MCI had higher levels of HbA1c and geriatric depression, and lower performance in all cognitive domains compared to non-MCI participants.

**Conclusion:** Although cognitive performance declined with age, HbA1c and geriatric depression played a greater role than age in predicting cognitive decline. This study highlights the prevalence of metabolism linked changes in cognition in community dwelling Indian older adults.

## 1. Introduction

It is well established that cognitive changes are a normal aspect of ageing. When decline in cognition exceeds an acceptable range of performance, it is typically a sign of a clinical or pathological condition. Understanding the pattern of cognitive change in aging individuals is important to distinguish normal aging from clinically impaired aging. At the same time, health conditions that influence cognition in aging also need to be understood. With the aging population increasing rapidly, globally as well as in India, understanding effects of aging and other related parameters on cognitive health is of utmost importance.

There are numerous studies on aging related cognitive changes in the North American and Western European populations. However, in the Indian context, there are only a handful of studies which have explored aging related cognitive change. Mathuranath and colleagues (2007) used Addenbrooke’s Cognitive Exam (ACE) and Mini Mental State Exam (MMSE) and found a linear relationship between cognitive performance and education in older adults (60-75yrs) from Trivandrum, Kerala. Another study on older adults (55-95yrs) in Kolkata, West Bengal, used a group of tests similar to those in the Montreal Cognitive Assessment (MoCA) and found decade-wise cognitive decline (Das et al., 2006). However, the authors did not describe whether the education levels were controlled before making decade-wise comparisons. Similarly, Tripathi and Tiwari (2011) observed that normally aging older adults with low levels of education were impaired on orientation and concentration using Hindi MMSE and a Brief Cognitive Rating Scale in urban older adults in Lucknow, Uttar Pradesh. However, 68% of their participants had completed only primary school or lower. A recent study profiled cognition using ACE and found that cognitive performance in middle-age and older-age adults (<70yrs) declined when compared to young adults, but no difference was observed between the middle-aged and older adults (Nigam and Kar, 2020).

Collectively, the tests used in the studies discussed so far are either MoCA, MMSE or ACE. While these tests are sensitive to detect dementia, they may not be sensitive to detect mild or early changes in cognition (Hugo and Ganguly, 2014). They do not provide an in-depth assessment of cognitive function which is necessary to confirm early changes in cognition (Petersen et al., 2004). Detailed cognitive assessments were carried out by Tripathi and colleagues (2014) on Indian older adults (55-64yrs) with education ranging from illiterates to college graduates to explore the effects of gender and education on cognitive function. The study observed that women performed better on memory tests and cognitive abilities were better in individuals with more years of education. However, the age-range of older adults included in the study was rather limited to generalize the observations.

Overall, studies from the Indian context give a screening level status of cognition function in the elderly. There is a need for detailed assessment of different cognitive domains to understand the trajectory of age-related cognitive changes in the Indian population.

Recent population surveys have shown high prevalence of diabetes in aging Indians (Jana and Chattopadhyay, 2022). Along with age, hyperglycemia may also affect cognitive performance. Associations of high glycated hemoglobin and cognitive change have also been observed in diverse populations (Jana et al., 2017; ‘Maan et al. 2021; Mimenza-Alvarado et al., 2020; Zheng et al., 2018). Recent studies in Indian adults (35-90yrs) have observed that high levels of HbA1c were linked to impaired performance on MoCA (Chakraborty et al., 2021; Lalithambika et al., 2019). These studies used screening level tools (MMSE, MoCA) to assess cognitive function which gives a limited understanding of changes in different cognitive domains. Few studies that assessed cognitive domains in-depth found deficits in executive function (Pappas et al., 2017), processing speed and working memory (meta-analysis, Mansur et al., 2018), and memory, language, attention, and visuo-spatial abilities (Dyer et al., 2021). A large UK cohort study by Antal and colleagues (2022) also observed deficits in executive function and processing speed in type-2 diabetes and found gray matter atrophy and altered brain activity across many brain areas, a sign of accelerated aging. The study, however, did not have measures of glycated hemoglobin (HbA1c) at the time of cognitive testing (Antal et al., 2022).

The role of vitamin B12 in modulating cognitive abilities is ambiguous. Some studies have shown that low vitamin B12 levels were associated low cognitive scores (Moorthy et al, 2012; Morris et al., 2012; Nalder et al., 2021), with a potential risk for Alzheimer’s disease (Lauer et al., 2022). However, many studies reported no association between vitamin B12 and cognition (Agnew-Blais et al., 2015; Zhang et al., 2020; Doets et al., 2013). Outcomes of interventions with vitamin B12 supplementation in older adults are also mixed. A few intervention studies show improved cognitive performance, especially when cognitive scores were low on MMSE (Sashindran et al., 2022) or in participants with mild cognitive impairment (de Jager et al., 2012). Some studies observed that vitamin B12, when supplemented with folic acid and vitamin B6 improved the serum homocysteine levels which may or may not translate to improvements in cognitive performance (Cheng et al., 2016; Dangour et al., 2015; Zhang et al., 2017). Other intervention studies found no association between vitamin B12 and cognitive performance in normally aging adults (Doets et al., 2013; Hvas et al., 2004, Markun et al., 2021).

Despite evidence of effects of aging and associations of hyperglycemia on general cognition, their differential contribution to cognitive changes in the Indian elderly population are not well established. Which domains of cognition are affected with increase in age? Are effects of hyperglycemia on executive function and processing speed also observed in Indian older adults? Is vitamin B12 linked to cognitive performance? In addition, we profiled participants for prevalence of mild neurocognitive disorder (mild NCD, DSM-5, 2022), also known as mild cognitive impairment (MCI) and explored whether glycated hemoglobin and vitamin B12 levels vary between participants profiled as MCI vs non-MCI. Age, glycated hemoglobin, or vitamin B12, which of these have a greater influence on cognitive performance? Through this study on Indian older adults, we sought answers to these questions.

## 2. Materials and Methods

### 2.1 Participants

This study was approved by the Institutional Ethics Committees at the National Center for Biological Sciences (NCBS) and the Institute of Ayurveda and Integrative Medicine – Foundation for Revitalization of Local Health Traditions (IAIM-FRLHT), both at Bengaluru, India. Participants were recruited through healthy aging awareness talks with a focus on cognitive health at elderly enrichment centers and citizen forums in Bengaluru. Participants were also recruited through snowball referrals (word of mouth). None of the participants had visited a clinician for any mental or cognitive health complaints. The participants themselves and their families considered them to be aging normally.

A total of 123 participants between 55-85 years completed this study. Participants were included if they had at least 10 years of school education and had basic English proficiency (read and communicate). Detailed health record and family history was taken. Participants were excluded if they had i) history of brain injury; ii) neurological or psychiatric illness; or iii) severe hearing or visual impairment. Details of participants are summarized in Table 1.

**Table 1.**
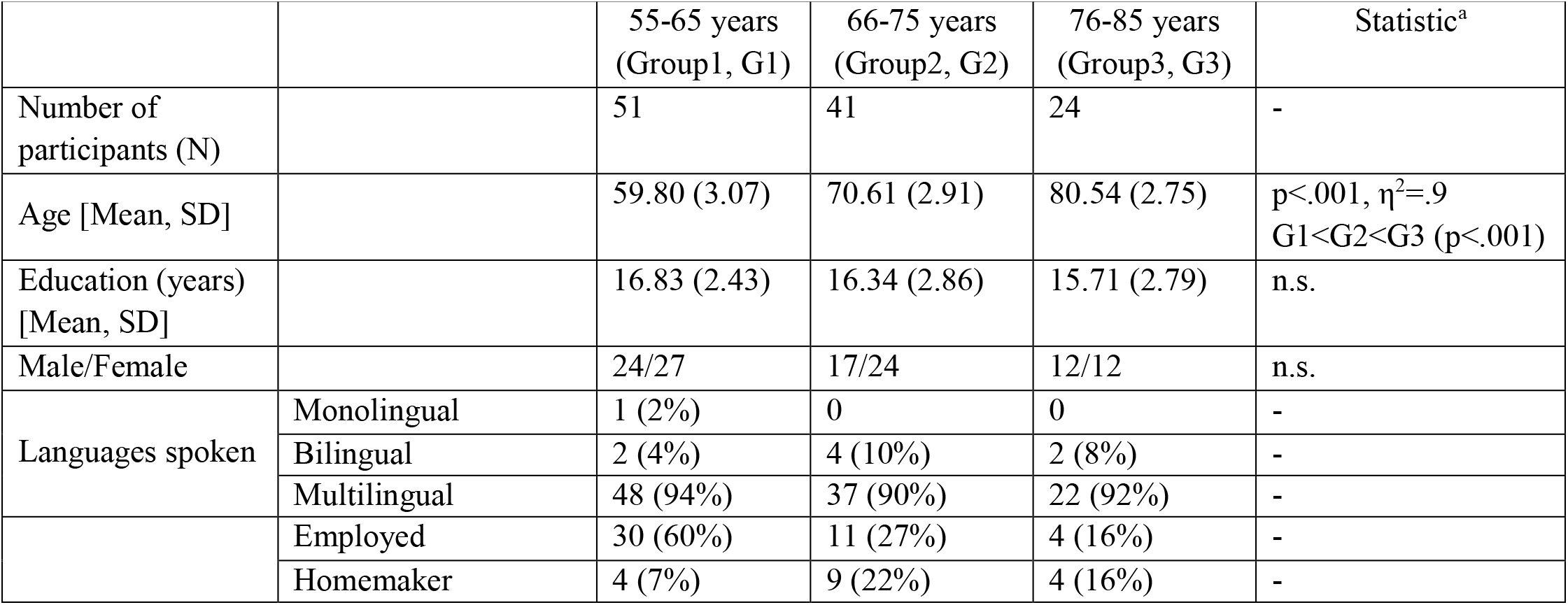
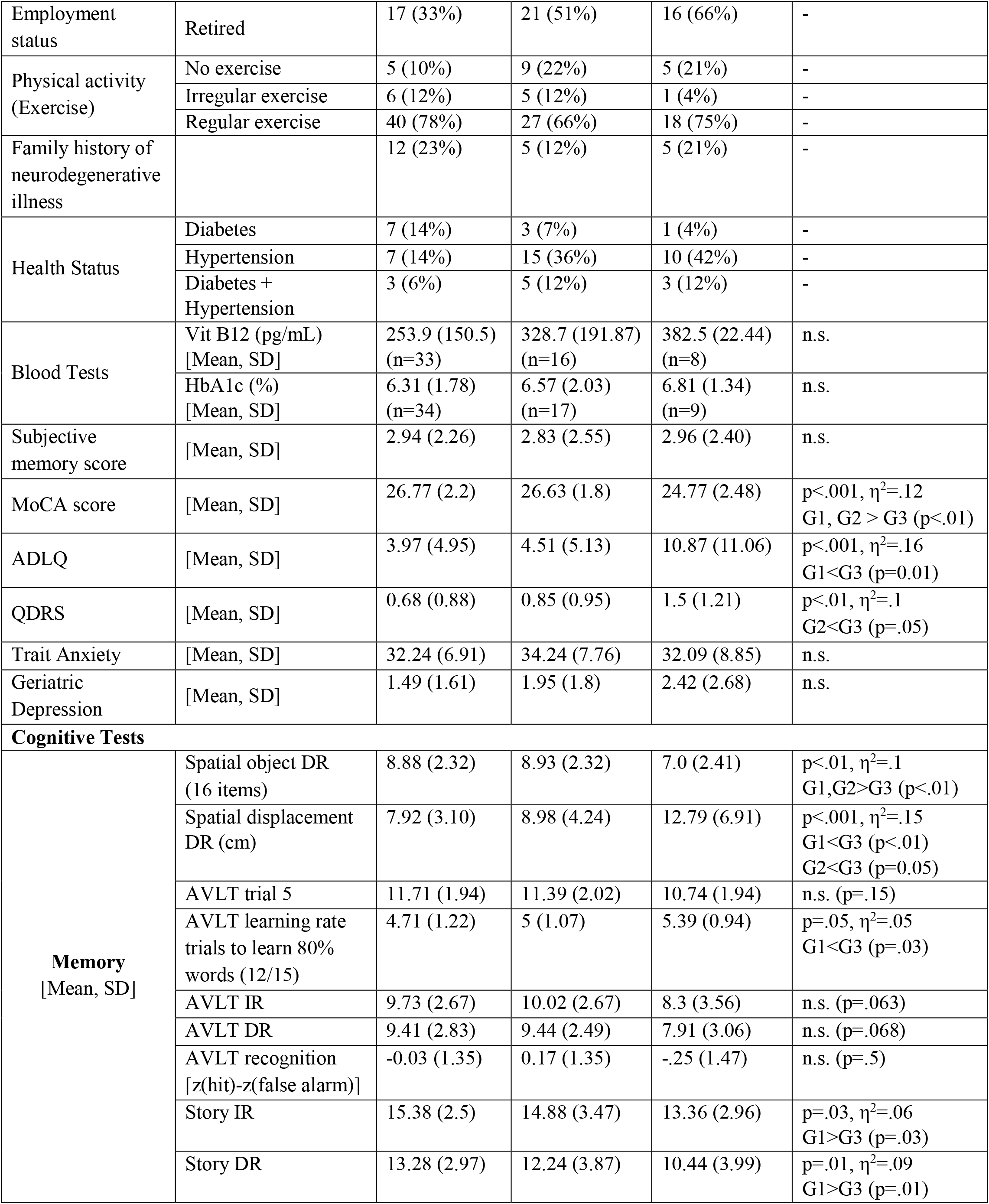
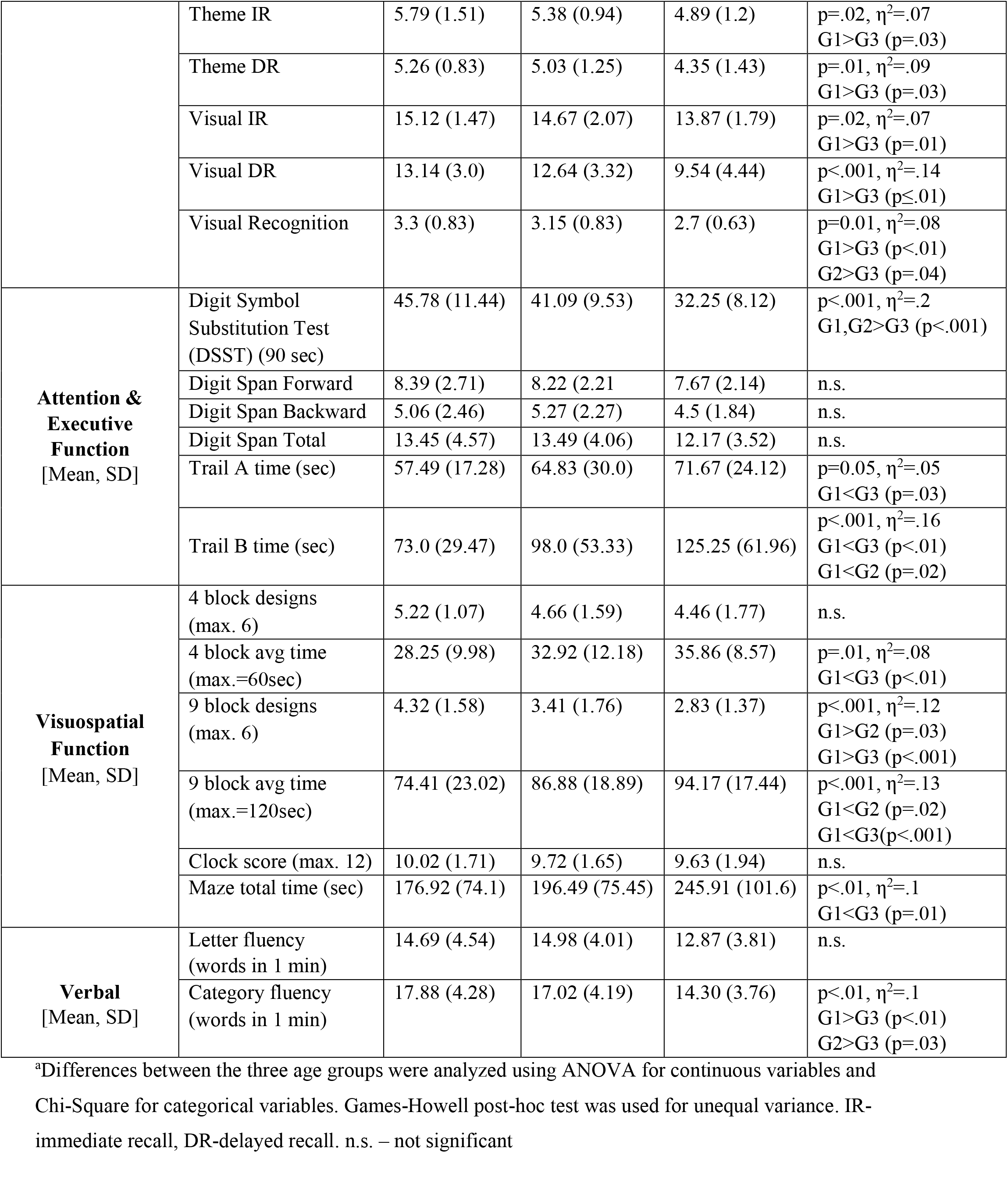
Summary of participant demographics and cognitive performance by age-decade.

### 2.2. Cognitive Assessments, Questionnaires, and Blood Parameters

All cognitive tests and questionnaires were administered in the paper-pencil format and were completely in 90-120 minutes.

#### 2.2.1. Measures to profile MCI

*Montreal Cognitive Assessment (MoCA)* has been shown to be sensitive in differentiating MCI from cognitively healthy participants (Chapman et al., 2016; Nasreddine et al. 2005; Roalf et al. 2013; Trzepacz et al., 2015). From a total MoCA score of 30, a score above 26 is considered normal and score below 19 is considered to be in the MCI range. It also accounts for less than 12 years of education by adding 1 point to the total score.

*Subjective Memory Complaint Questionnaire* developed by Youn and colleagues (2009) was used. Participants responded “yes” or “no” to 14 questions that cover everyday, spatial, global, and prospective memory. These questions helped to prod older adults to review themselves on different memory functions.

*Activities of Daily Living (ADLQ)* is a questionnaire filled by informants (spouse/child) (Johnson et al., 2004). It is sensitive to mild decline and measures functional abilities in 6 areas - self-care, household care, employment and recreation, shopping and money, travel, and communication. The scoring gives a gradation of impairment of functional abilities (mild 0-33%, moderate 34 - 66%, and severe > 67%). ADLQ correlates inversely with MoCA (Durant et al., 2016). To include changes due to normal aging, individuals with ADLQ scores only less than 33% were included.

*Quick Dementia Rating Scale (QDRS)* comprises of 10 questions related to cognitive, behavioral, and functional abilities. The first six questions in the QDRS give the Clinical Dementia Rating (CDR) Scale equivalent score (Galvin, 2015). The informant (spouse/child) has to compare the participant’s past-present abilities. The key feature is the change in cognitive abilities. The total score ranges between 0-30. Participants with scores ≥ 5, indicative of early dementia, were excluded. This scale is shorter than CDR and efficient for screening community members for research purposes (Berman et al., 2017).

*Geriatric Depression Scale (GDS)* is a screening tool for depressive symptoms in older adults (Sheikh and Yesavage, 1986). It has 15 questions with a ‘yes’ or ‘no’ response. Participants have to rate their feelings over the past one week. Scores above 10 are indicative of depression (Greenberg, 2007).

*Spielberger’s Trait Anxiety Inventory (STAI)* is a 20-item scale to evaluate levels of anxiety generally experienced by an individual, with scores ranging between 20-80 (Spielberger et al., 1983). This was collected to observe anxiety levels in an older Indian population.

#### 2.2.2. Cognitive tests

##### A. Memory tests

*Spatial Memory Test* was developed by Smith and Milner (1981) to test memory for objects and their spatial locations. A set of 16 toy objects were randomly placed on a 60cm x 60cm sheet of paper before the participant came into the room. Toy objects were doll-sized and included everyday items (bowl, bucket, car, chair, computer, cup, dress, hairbrush, necklace, pot, scissors, shoes, spanner, spatula, telephone, umbrella). For encoding, participants had to name each toy object and estimate its price if it were a real item. Objects were then taken out of sight and kept aside for a 90 min interval that was filled by other cognitive tests. At the end of the interval, for recall, participants had to name as many objects as they could remember. Subsequently, participants were given all the objects and were instructed to place them in their original locations on the paper. The original and recalled spatial positions of the objects were photographed with a top view of the sheet and were overlaid to calculate the absolute displacement (cm) of each object using ImageJ, an open-source software for analyzing images.

*Auditory Verbal Learning Test* (AVLT) assessed auditory verbal learning and memory. The test comprised of 15 unrelated words (List A) that were read aloud, one word per second, followed by a free recall. This was repeated over 5 trials. Subsequently, another list of 15 unrelated words (List B) was read aloud followed by a free recall. List B was read only once and served as interference (distractor list), followed by free recall of List A words (immediate recall). List A was recalled again after 20-30 mins (delayed recall), followed by a recognition of List A words. The recognition test included words from List A, List B, 15 semantically related words to List A, and 15 phonetically related words to List A. The standardized difference between hits and false alarms was the recognition score.

*Logical (Story) Test* consisted of listening and remembering a short story. Two passages from the Wechsler Memory Scale-III (WMS-III INDIA; Wechsler & Gurappa, 2009) adapted for the Indian population were used. Each passage was read aloud slowly and clearly in a neutral tone followed by an immediate and delayed recall (20-30 min). Each passage included 25 facts (details) and 7 themes (gist). Participant recall was audio-recorded and scored later.

*Visual Reproduction Test* assessed visual memory. Four figures were taken from the Wechsler Memory Scale-III (WMS-III INDIA; Wechsler & Gurappa, 2009). Participants were given 10 seconds to observe each stimulus figure and then taken out of sight. Then a blank sheet of paper was given and participants were instructed to draw the observed figure (immediate recall). Delayed recall for all 4 figures was conducted after 20-30mins followed by a recognition test, which included the original figure and 5 slightly distorted versions shown together. The participant had to identify the original figure. For scoring, the figures were analyzed by different shapes and their relative location in the figure. The point-scoring format was taken from the Wechsler Memory Scale.

##### B. Attention and Executive Function

*Digit Symbol Substitution Test* measured information processing and motor speeds, taken from the Wechsler Adult Intelligence Scale—Fourth Edition (WAIS–IV; Wechsler, 2008a). Participants had to match numbers to symbols. Number-symbol pairs were presented on top of a sheet and numbers from 1-9 in random order were printed below in multiple rows. Below each number participants had to draw the corresponding symbol as quickly as they could. The total number of correctly drawn symbols within 90 seconds constituted the score.

*Digit Span Test* measured attention and working memory. Random sequences of numbers were read aloud at the rate of 1 digit per second. Participants had to listen carefully and repeat the numbers in the same order (forward span). The number of digits in the sequence increased serially. The listen-repeat protocol continued until participants made errors on two consecutive sequences. The same protocol was then used for backward span where participants had to repeat the numbers in the reverse order. The total number of correctly repeated forward and backward sequences comprised the score.

*Trail Making A&B Test* had two parts - Trail A primarily reflects visuo-motor abilities while Trail B measures working memory and attention shifting. In Trail A, participants had to connect circles numbered 1-25 presented in random locations on a sheet of paper. In Trail B, participants had to alternate between numbers (1-12) and letters (A-K) while connecting the dots (1-A-2-B). Time taken to complete both trails was noted.

##### C. Visuospatial Abilities

*Block Design Test* measured visuo-spatial abilities and was taken from the Weschler Memory Scale, 3^rd^ Indian edition. Participants were shown abstract designs which had to be reconstructed by assembling red and white blocks. Each of the 4-block designs (n=6) had to completed within 60 seconds and 9-block designs (n=6) withing 120 seconds. The time taken to assemble the designs correctly was noted. If the design was not completed in the given time, the number of correctly placed blocks within the time limit were noted.

*Maze Test* can identify deficits in route planning and foresight in older adults (Snellgrove, 2005; Staplin et al., 2013). Two standard mazes (no. 12 and no. 14) were taken from the Porteus Tests (Porteus, 1933). The Snellgrove maze was also included. The time taken to complete the mazes was noted. Dead-end entries were scored as errors.

*Clock Orientation Test* measured mental rotation and egocentric spatial processing (Coughlan et al. 2018). Participants had to imagine that they were standing in the center of a large clock and facing a particular number (e.g. 12). They were asked to point to the direction of different numbers (e.g. 3 – right). The orientation becomes increasingly difficult over 12 questions. Time taken to respond and accuracy was noted.

##### D. Verbal (Language) Abilities

*Letter Fluency* assessed phonemic fluency. Participants were instructed to generate words starting from a given letter within one minute, excluding names of people and places.

*Category Fluency* included naming as many animals as possible within one minute.

#### 2.2.3. Testing of Blood Parameters

A subset of the participants (n=60) participated further in another leg of the study and provided blood samples. A trained phlebotomist visited the homes of consenting participants. Blood was collected 2-3 hours post-prandial. Samples were taken by the phlebotomist for analysis to a private lab certified by the National Accreditation Board for Testing and Calibration Laboratories (NABL), India. Blood parameters included in this study are glycated hemoglobin (HbA1c, %) and vitamin B12 (pg/ml). Glycated hemoglobin (HbA1c) is a commonly used diagnostic marker of presence and severity of type 2 diabetes.

## 3. Data collection time line and procedure

Interested participants were given a choice to visit the research center or have a researcher visit their home to administer the cognitive tests. Participants were briefed about the purpose and duration of the study. After obtaining the written consent and providing general instructions, demographic details and health history were collected. The spatial memory test was always administered first in order to accommodate the 60–90minute time interval between encoding and recall. All other cognitive tests were administered randomly. Intervals of delayed recall were filled with other cognitive tests. Blood samples were collected within a week of cognitive testing.

## 4. Results

Data from 116 participants is included in the analysis. Three participants with QDRS ≥ 5 and four participants with geriatric depression score >10 were excluded from the analysis (Figure 1). Table 1 summarizes participants’ demographics of included participants.

**Figure 1.**
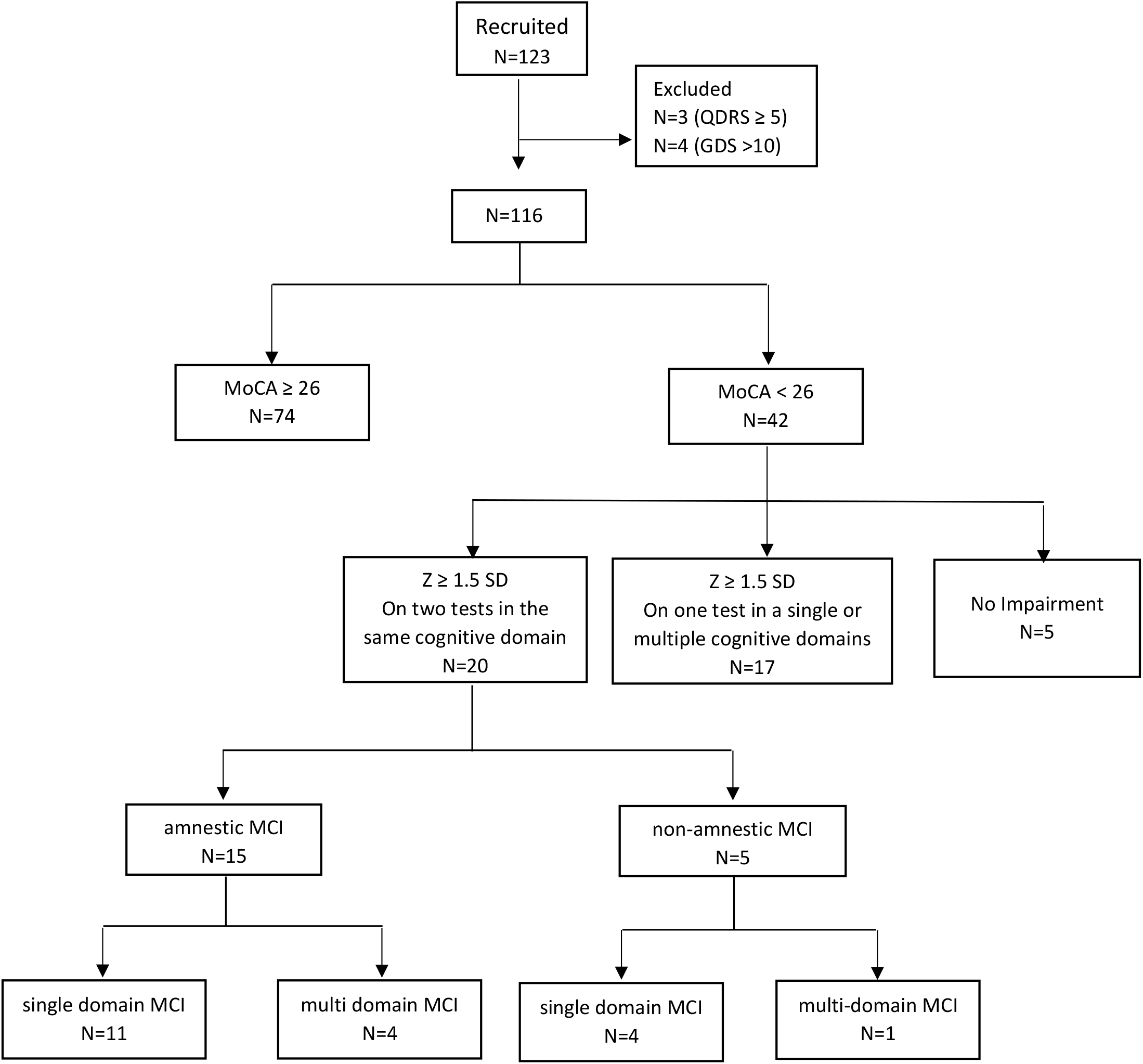
Classification of participants with MoCA<26 into sub-types of MCI. Z=normalized standard deviation.

### Classification of cognitive status

The cognitive status of the participants was determined using MoCA and cognitive tests covering different cognitive domains. Participants with MoCA≥26 were divided into 3 age groups (55-65, 66-75, 76-85) and their cognitive performance was used as age-appropriate cognitive norms to calculate the extent of cognitive impairment. MCI was determined using four-point criteria, namely i) MoCA<26; ii) reduction in performance by ≥ 1.5 normalized standard deviations on two or more tests in the same or multiple cognitive domains when compared to average performance of age decade-matched participants with MoCA ≥ 26; iii) daily living activities unaffected (ADLQ <33%); iv) No dementia (QDRS < 5) (Sósa et al., 2012; Petersen, 2004). Since this study included community dwellers, subjective complaint was not included as criteria to confirm MCI. Participants with MoCA<26 but no impairment (<1.5 normalized standard deviation on all cognitive tests) were considered non-impaired (Dautzenberg et al, 2020). Figure 1 illustrates the classification of participants with MoCA<26 in to sub-types of MCI. Analysis using the Mann Whitney U test showed that performance on most cognitive test was lower in the MCI group (MoCA<26, n=20, ref Fig 1) compared to normal group (MoCA≥26, n=74). The results are summarized in Table 2.

**Table 2.**
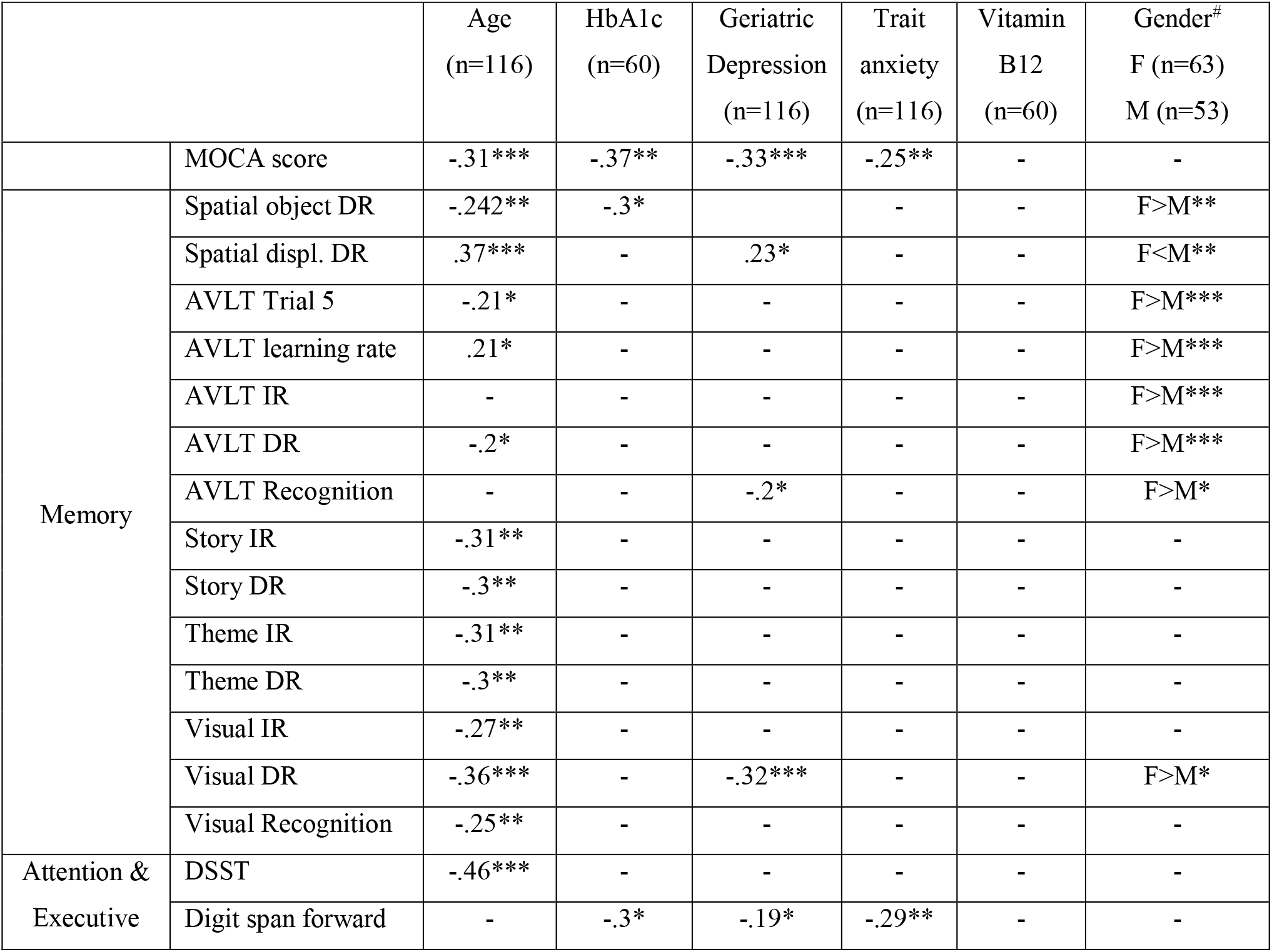
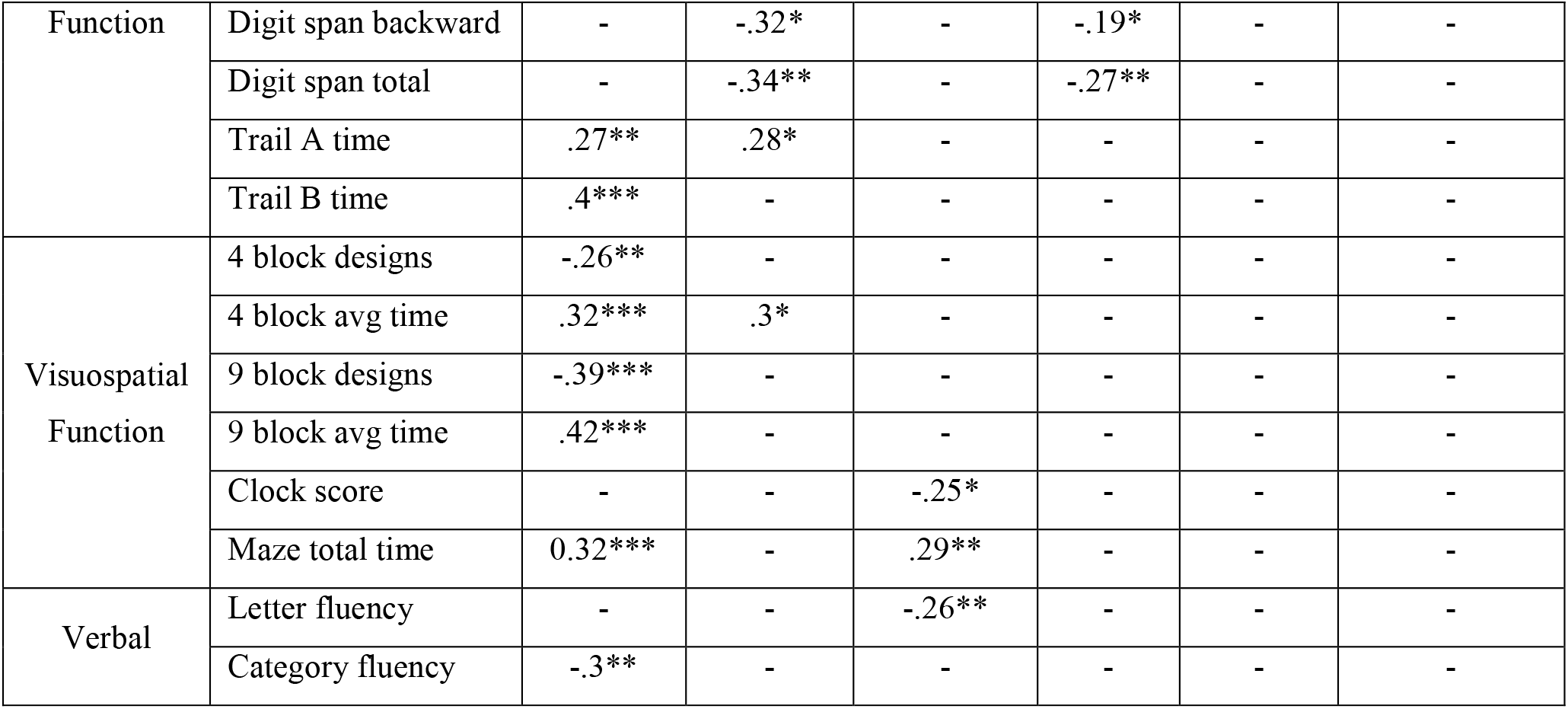
Significant correlations between cognitive tests and various parameters. *p<0.05, **p<0.01, ***p<0.001. # T-test

### Age

The Pearson correlation showed a significant negative association between age and MoCA scores, r= - 0.32, n=116, p<.001 (Figure 2A). An effect of age on MoCA performance was observed using a one-way ANOVA (F_(2,113)_ = 7.91, MSE= 4.56, p<.001, η_p_^2^= .12), Figure 2B. Games-Howell post-hoc test for unequal variances showed that the 76-85 age group had significantly lower MOCA scores than 66-75 age group (p=.007) and 55-65 age group (p=.005). There was no significant difference in MoCA scores between 55-65 and 66-75 age groups (p=.9). Effects of age by decade on performance on all cognitive tests is summarized in Table 1. Pearson correlations of age with performance on all cognitive tests are summarized in Table 2.

**Figure 2.**
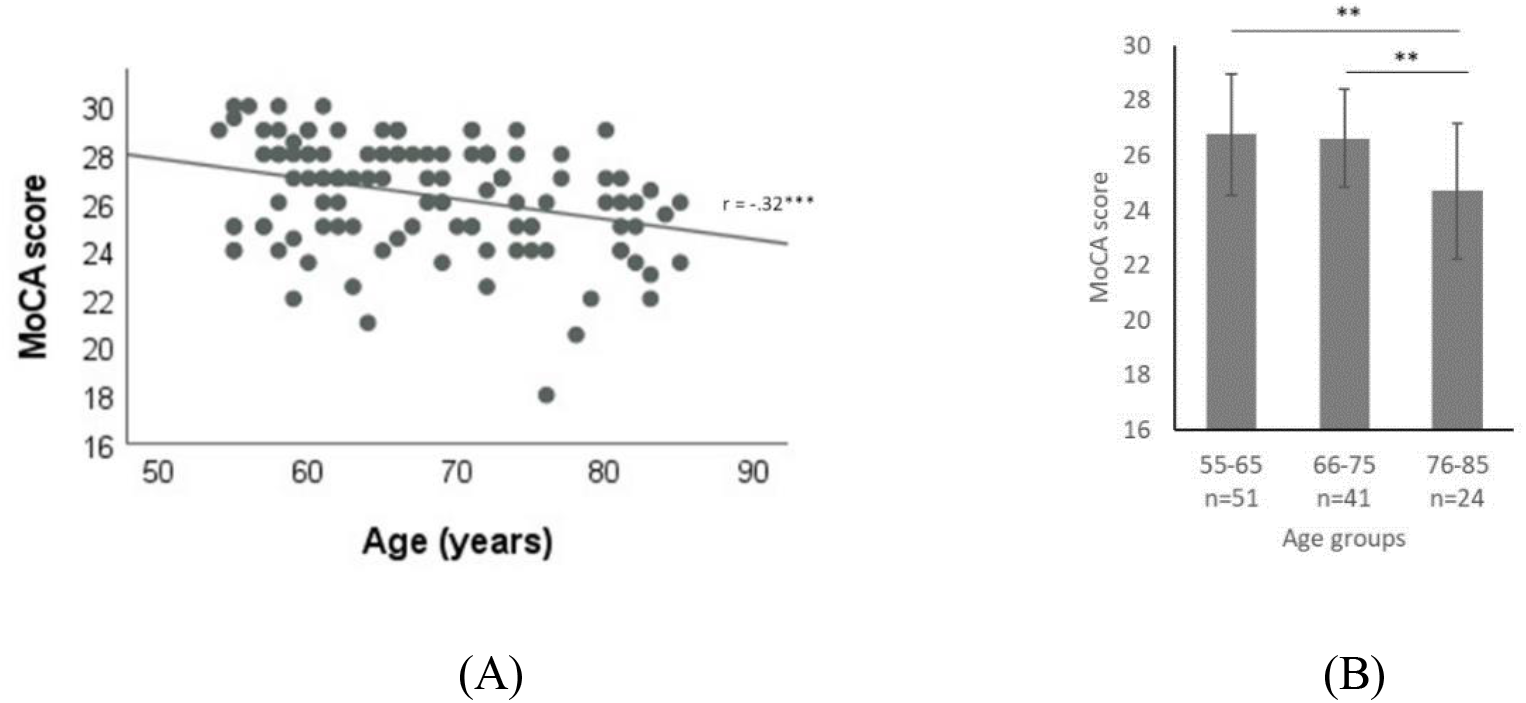
A) Scatterplot of negative correlation between age and MoCA scores. B) Bar-graph of MoCA scores participants by age decade. Error bars denote standard deviation. ***p<0.001, **p<0.01.

### Glycated Hemoglobin (HbA1c)

Pearson correlation analysis showed that HbA1c levels and MoCA scores were significantly negatively correlated (r= -.37, n=60, p=.004). HbA1c levels also correlated negatively on tests of spatial object recall (r=.3, p=.02) and digit forward span (r= -.3, p=.02), backward span (r= -.32, p=.01), and total span (r= - .34, p=.008), as summarized in Table 2. HbA1c levels correlated positively with completion times for Trail A (r=.28, p=.03) and 4 block design (r=.3, p=.02), summarized in Table 2. HbA1c values were split into two groups, normal level (<7%, n=50) and high level (≥7%, n=10). A Mann Whitney U test compared the normal and high HbA1c groups and found significant differences in MoCA scores (U=119, p=.009, r =.34), digit forward span (U=131, p=.017, r =.31), backward span (U=114.5, p=.007, r =.35), total span (U=103.5, p=.004, r =.38), DSST (U=142, p=.032, r =.28), and completion time for Trail A (U=155, p=.05, r=.24) Trail B (U=109.5, p=.005, r =.36), 4-blocks (U=113.5, p=.007, r=.35), and 9-blocks (U=130.5, p=.018, r=.31). These effects are summarized in Figure 3. MCI participants (MoCA<26, n=20, ref Fig.1) had higher HbA1c values compared to normal participants (MoCA≥26, n=74) on a Mann Whitney U test (U=129.5, p=.05, r=.27) (Table 3).

**Table 3.**
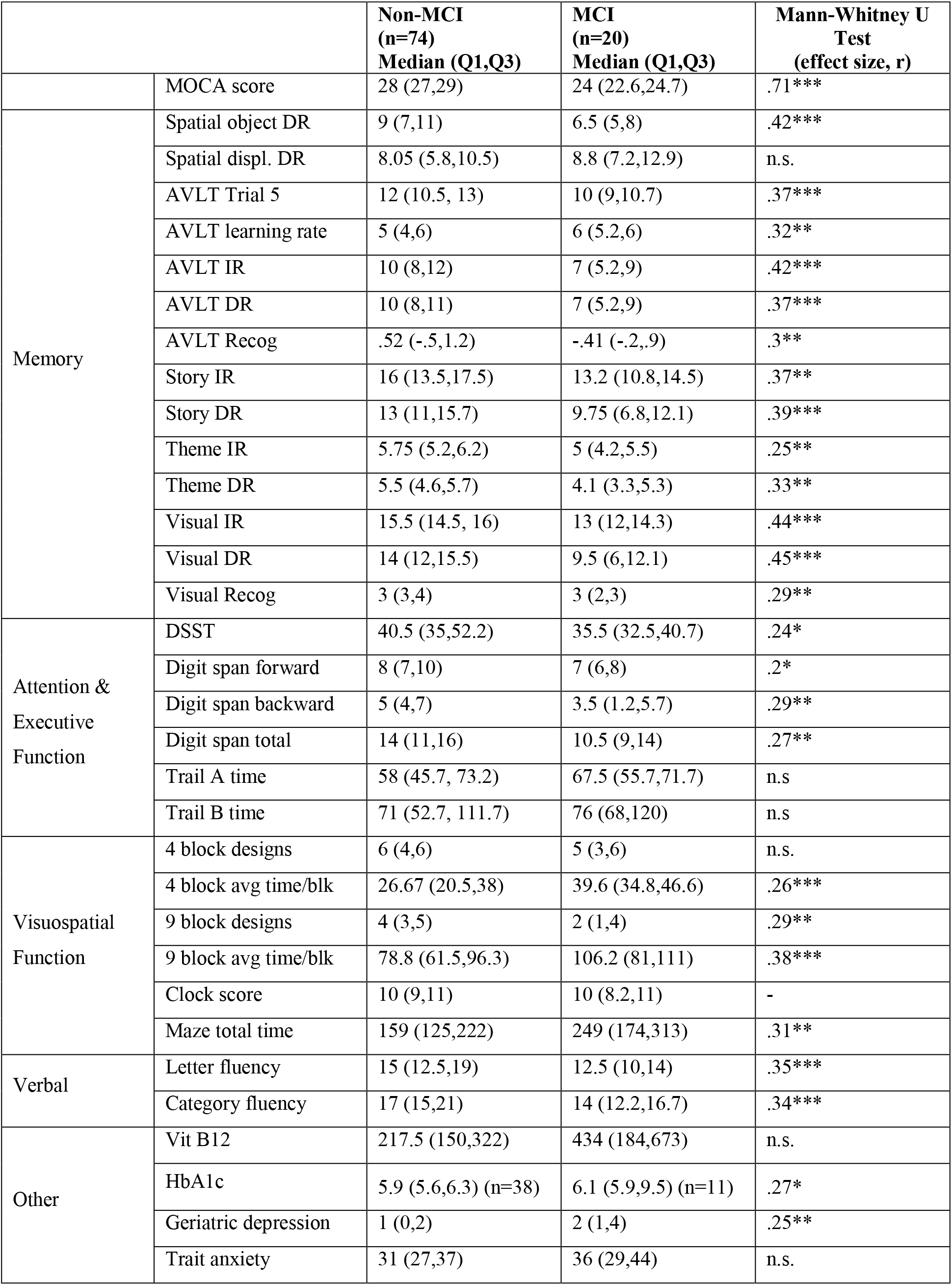
Non-MCI vs MCI differences in cognitive performance. *p<0.05, **p<0.01, ***p<0.001.

|  |  | **Non-MCI**<br>**(n=74)**<br>**Median (Q1,Q3)** | **MCI**<br>**(n=20)**<br>**Median (Q1,Q3)** | **Mann-Whitney U**<br>**Test**<br>**(effect size, r)** |
| --- | --- | --- | --- | --- |
|  | MOCA score | 28 (27,29) | 24 (22.6,24.7) | .71*** |
| Memory | Spatial object DR | 9 (7,11) | 6.5 (5,8) | .42*** |
|  | Spatial displ. DR | 8.05 (5.8,10.5) | 8.8 (7.2,12.9) | n.s. |
|  | AVLT Trial 5 | 12 (10.5, 13) | 10 (9,10.7) | .37*** |
|  | AVLT learning rate | 5 (4,6) | 6 (5.2,6) | .32** |
|  | AVLT IR | 10 (8,12) | 7 (5.2,9) | .42*** |
|  | AVLT DR | 10 (8,11) | 7 (5.2,9) | .37*** |
|  | AVLT Recog | .52 (-.5,1.2) | -.41 (-.2,.9) | .3** |
|  | Story IR | 16 (13.5,17.5) | 13.2 (10.8,14.5) | .37** |
|  | Story DR | 13 (11,15.7) | 9.75 (6.8,12.1) | .39*** |
|  | Theme IR | 5.75 (5.2,6.2) | 5 (4.2,5.5) | .25** |
|  | Theme DR | 5.5 (4.6,5.7) | 4.1 (3.3,5.3) | .33** |
|  | Visual IR | 15.5 (14.5, 16) | 13 (12,14.3) | .44*** |
|  | Visual DR | 14 (12,15.5) | 9.5 (6,12.1) | .45*** |
|  | Visual Recog | 3 (3,4) | 3 (2,3) | .29** |
| Attention &<br>Executive<br>Function | DSST | 40.5 (35,52.2) | 35.5 (32.5,40.7) | .24* |
|  | Digit span forward | 8 (7,10) | 7 (6,8) | .2* |
|  | Digit span backward | 5 (4,7) | 3.5 (1.2,5.7) | .29** |
|  | Digit span total | 14 (11,16) | 10.5 (9,14) | .27** |
|  | Trail A time | 58 (45.7, 73.2) | 67.5 (55.7,71.7) | n.s |
|  | Trail B time | 71 (52.7, 111.7) | 76 (68,120) | n.s |
| Visuospatial<br>Function | 4 block designs | 6 (4,6) | 5 (3,6) | n.s. |
|  | 4 block avg time/blk | 26.67 (20.5,38) | 39.6 (34.8,46.6) | .26*** |
|  | 9 block designs | 4 (3,5) | 2 (1,4) | .29** |
|  | 9 block avg time/blk | 78.8 (61.5,96.3) | 106.2 (81,111) | .38*** |
|  | Clock score | 10 (9,11) | 10 (8.2,11) | - |
|  | Maze total time | 159 (125,222) | 249 (174,313) | .31** |
| Verbal | Letter fluency | 15 (12.5,19) | 12.5 (10,14) | .35*** |
|  | Category fluency | 17 (15,21) | 14 (12.2,16.7) | .34*** |
| Other | Vit B12 | 217.5 (150,322) | 434 (184,673) | n.s. |
|  | HbA1c | 5.9 (5.6,6.3) (n=38) | 6.1 (5.9,9.5) (n=11) | .27* |
|  | Geriatric depression | 1 (0,2) | 2 (1,4) | .25** |
|  | Trait anxiety | 31 (27,37) | 36 (29,44) | n.s. |

**Figure 3.**
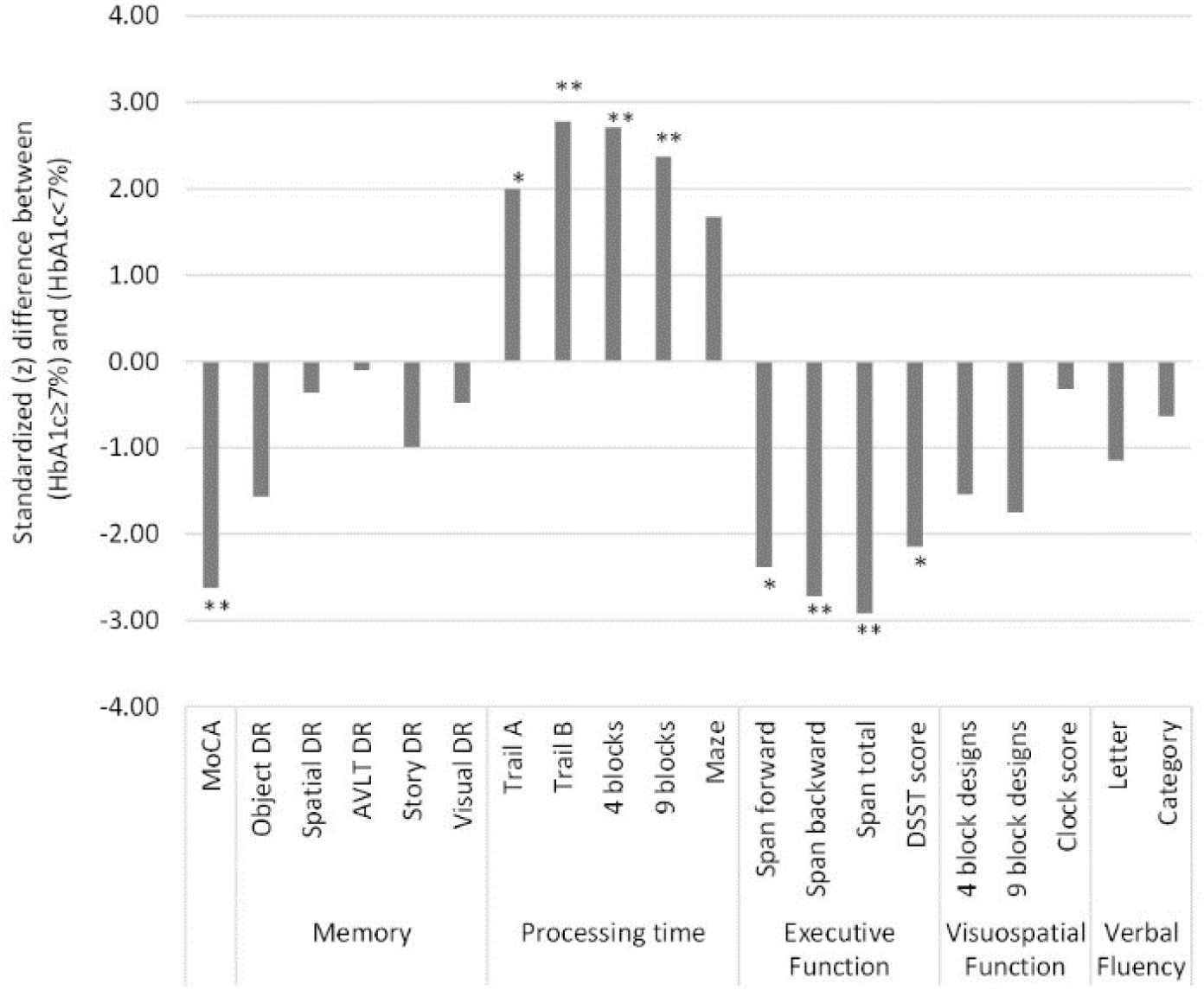
Standardized (Z) difference between participants with HbA1c≥7% (n=10) and HbA1c < 7% (n=50). Z scores from Mann-Whitney U test. **p<0.01, *p<0.05.

### Vitamin B12

Pearson correlations between vitamin B12 level and individual cognitive test scores revealed no significant associations. Since vitamin B12 levels between 200-900 pg/ml are considered normal (Wolters et al., 2004), participants were split in low vitamin B12 (<200, n=25) and normal (≥200, n=35) groups. Mann Whitney U test showed no difference between low and normal vitamin B12 groups on MoCA scores (U=285, p=.18) or any other cognitive test. Conversely, MCI and non-MCI participants also showed no difference in vitamin B12 levels on Mann Whitney U test (U=95, p=.06).

### Psychosocial factors

#### Geriatric depression

Pearson correlations revealed that geriatric depression scores correlated with MoCA scores (r= -.33, p<.001), average spatial displacement (r=.23, p=.014), visual delayed recall (r= -.32, p<.001),, letter fluency (r= -2.6, p=.004), maze completion time (r=.29, p=.002), Clock test (r= -.25, p=.013), summarized in Table 2. Levels of geriatric depression were higher in MCI participants than normal participants on a Mann Whitney U test (U=479, p=.013, r=.25) (Table 3).

#### Trait anxiety

Pearson correlations showed that trait anxiety correlated negatively with MoCA scores (r=-.25, p=.007), digit span forward (r=-.29, p=.002), digit span backward (r=-.19, p=.043), total digit span (r=-.27, p=.004), (Table 2). There was no difference in trait anxiety scores between MCI and non-MCI participants.

#### Gender

Effect of gender on cognitive performance analyzed using a Mann Whitney U test showed than females performed better than males only on tests of memory – spatial object recall (U=1191.5, p=.008, r=.25), average spatial displacement (U=1176, p=.009, r=.24), number of AVLT words learnt by Trial 5 (U=768, p<.001,r=.46), AVLT learning rate (trials taken to learn 12 words/80%) (U=934.5, p<.001, r=.4), AVLT IR (U=836.5, p<.001, r=.42), AVLT DR (U=847, p<.001, r=.42), AVLT Recognition (U=1118.5, p=.01, r=.24), visual DR (U=1171.5, p=.018, r=.22), summarized in Table 2.

#### Multiple regression of factors (age, HbA1c, geriatric depression, trait anxiety) on MoCA

Taking the factors that correlated significantly with MoCA, a stepwise multiple regression analysis was carried out to check if age, HbA1c, geriatric depression or trait anxiety could predict MoCA score. The regression analysis indicated that 2 of the predictors – geriatric depression (β= -.41, p<.001) and HbA1c (β= -.33, p=.004) contributed significantly to 28% of the variance in MoCA score (adjusted R^2^ = .28, F_(2,57)_ = 12.59, p<.001) (Table 4). Age and trait anxiety did not reach statistical significance. Semi-partial correlations revealed unique contributions of geriatric depression (17%) and HbA1c (11%) to the 28% variance in MOCA score (Figure 4).

**Table 4.** Stepwise multiple regression of age, HbA1c, geriatric depression, and trait anxiety

| | B | 95% CI (B) | $\beta$ | $r_{sp}^2$ | p-value | Adjusted $R^2$ |
| --- | --- | --- | --- | --- | --- | --- |
| Constant | 30.64 | [28.4, 32.81] |  |  |  |  |
| Geriatric depression | -0.49*** | [-0.75, -0.23] | -0.41 | 0.17 | <.001 | .28 |
| HbA1c | -0.48** | [-0.8, -0.16] | -0.33 | 0.108 | .004 |  |
| *<u>Variables rejected</u>* |  |  |  |  |  |  |
| Age | -.07 |  | -.23 |  | .06 |  |
| Trait anxiety | -.03 |  | -.07 |  | .61 |  |

B – unstandardized coefficient, $\beta$ – standardized coefficient, $r_{sp}^2$ – semi-partial correlation squared

**Figure 4.**
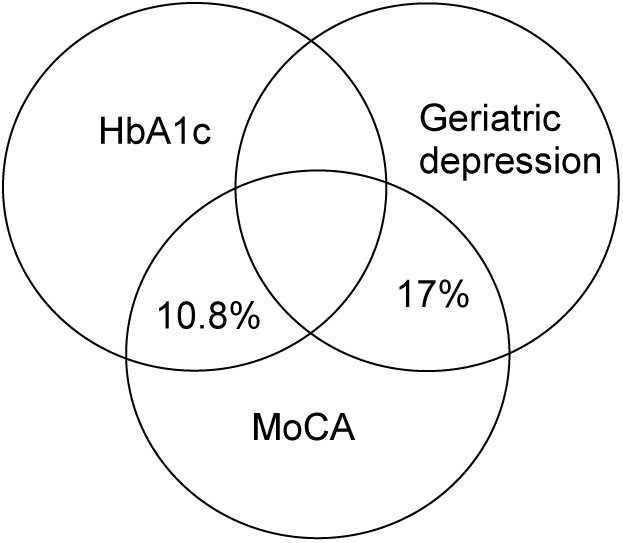
Unique contribution of factors (HbA1c, geriatric depression) in the variance observed in MoCA scores.

## 5. Discussion

Considering the rising aging population along with high incidence of diabetes in India, cognitive aging and its relationship to blood glucose is not well understood in the Indian population. To fill this gap, we explored the effect of age and associations of glycated hemoglobin, vitamin B12, geriatric depression, and anxiety on performance in different cognitive domains in community dwelling urban Indian older adults. In the following sections, we discuss the influence of each factor.

### Age

We observed that with increasing age MoCA scores (Figure 2A) and performance in all cognitive domains gradually declined (Table 2). However, the difference in performance was statistically significant only between 55-65yr and 76-85yr groups (Table 1 & Figure 2B). It appears as though the decline becomes apparent approximately around the age of 70 in the aging population. This trend was also noted by Tripathi & Tiwari (2011). Additional recent evidence suggests that sudden cellular and molecular decline occurs after the age of 70 which may translate to cognitive decline (Mitchell et al., 2022), which further supports our observations.

Here, we also discuss performance on cognitive tests where effects of age are not straightforward. In AVLT, although delayed recall gradually declined with age (Table 2), the difference in delayed recall across age decades did not reach statistical significance (Table 1). Repetition over 5 trials of learning may have helped participants to retain words well compared to other memory tests, where participants are exposed to the stimuli only once. However, the rate of learning declined with age, participants took more trials to learn the items as age increased (Table 1 and 2). We also found that females performed better than males on the AVLT, an effect that has also been observed by others in the Indian (Tripathi et al., 2014), Israeli (Vakil & Blachstein, 2007), Australian (Geffen et al., 1997), and Chinese (Guo et al., 1993) populations.

Letter fluency was unaffected by age while category fluency declined with age. Other studies have also noted such a dissociation and have shown that letter fluency may be affected by education whereas category fluency may be affected by age (Tombaugh et al., 1999; Cuadrado et al., 2002; Elgamal et al., 2011; Mathuranath et al., 2003). In this study education levels across decade-wise age groups were similar and no effects of age were seen on letter fluency.

Age also had no effect on forward or backward digit span performance. Since the study group consisted of only older adults, the effects on digit span across each decade of older life may be minimal. Similar null effects of age on digit span were reported in another study on Indian older adults (Tripathi et al., 2014) and in a Hispanic cohort (Pontón et al., 1996), where education correlated positively with digit span performance. Tripathi and colleagues (2019) have also reported a large effect of literacy on forward and backward span in an urban Indian cohort. Upon analyzing, we too found that years of education correlated positively with digit ***forward*** span performance (r=.26, p=.005), a small effect since the majority of our participants had higher levels of education.

We observed no effect of age on clock test performance. The clock test was designed to capture effects of aging on egocentric spatial processing (Coughlan et al., 2018). However, more recent studies have shown that egocentric spatial processing is relatively less impaired in older adults compared to allocentric spatial processing (Colombo et al., 2017; McAvan et al., 2021).

Overall, a gradual decline in cognitive abilities was observed as age advanced with noticeable differences post-70 years. Years of education played a protective role on tests of verbal fluency and executive function (digit span), where performance remained unaffected by age.

### Glycated Hemoglobin (HbA1c)

Participants with higher levels of HbA1c had lower scores on MoCA, took longer to complete trail making and block design tasks indicating slower processing time, and were impaired on digit span and digit symbol substitution (executive function) (Figure 3). Executive function and processing speed are dependent on the frontal regions that are typically affected by glucose dysregulation (Hishikawa et al., 2014; Weinstein et al., 2015). These results align with those observed in other cohorts (Mansur et al., 2018; Pappas et al., 2017). Performance on tests of memory, visuospatial abilities, and verbal fluency were unaffected by HbA1c levels. Others have also reported similar dissociations, where executive function and processing speed were affected by high glucose levels but memory remained unaffected (Antal et al., 2022; Casagrande et al., 2021; Nandipati et al., 2012). Many of our participants were on anti-diabetic medication which has been observed to rescue memory impairments (Alagiakrishnan et al., 2013; Sritawan et al., 2021; Zhong et al., 2018). Such medication might have a role in protecting the decline in memory in our cohort.

We also observed a non-significant but trending difference in HbA1c levels and MoCA scores due to exercise. No-exercise participants has higher HbA1c levels and lower MoCA scores (n=10, HbA1c = 7.6%, MoCA = 25.6) than regular exercise participants (n=45, HbA1c = 6.2%, MoCA = 26.5). Studies in elderly clinical populations have observed beneficial effects of exercise on memory in MCI participants (Suzuki et al., 2013) and HbA1c levels in diabetic participants (Castaneda et al., 2002). However, further studies are needed to understand the relationship between exercise, HbA1c, and cognitive function in non-clinical populations.

Individuals with impaired glucose metabolism not only show decline in cognition but can also develop white matter hyperintensities (Hishikawa et al., 2014; Weinstein et al., 2015). Such vascular abnormalities are a risk for vascular dementia (Ott et al., 1996; Raffaitin et al., 2009; Vogelgsang et al., 2018). Community dwelling participants in our study that were profiled as MCI had higher levels of HbA1c than non-MCI participants (Table 3) which highlights the prevalence of metabolism linked changes in cognition in a non-clinical population who may as well be at risk for vascular dementia.

### Vitamin B12

There were no associations between vitamin B12 levels and MoCA scores or any other cognitive score. Similar outcomes have been noted in other studies (Agnew-Blais et al., 2015; Zhang et al., 2020; Doets et al., 2013). There appears to be a complex relationship between vitamin B12, other vitamins, and folic acid in regulating serum homocysteine levels. Folic acid regulates vitamin B12 levels, and when vitamin B12 levels drop, serum homocysteine levels go up which may lead to neurodegeneration (Morris et al., 2007). As most of our participants were in the normal but lower range of vitamin B12, the effect of varying levels of vitamin B12 could not be observed.

### Geriatric Depression

Geriatric depression scores of included participants were negatively associated with performance on visuo-spatial tasks (spatial memory, delayed visual reproduction, maze completion time, clock score). Visuospatial tasks are dependent on the hippocampus, a region in the medial temporal lobe, whose function is modulated by cholinergic and serotonergic activity (Ridley et al.,1988). The activity of these neurotransmitters is sensitive to geriatric depressive modulation (Nobler et al., 1999) and may alter hippocampal activity (Gunning and Smith, 2011). Similar observations of late life depression affecting visuospatial performance have been reported in a Slovenian elderly population (Klojčnik et al., 2017).

Interestingly, the participants profiled as MCI had higher geriatric depression scores non-MCI participants, even after the excluding geriatric depression scores >10 from the analysis. Similarly, another study excluded clinically depressed older adults and yet found higher depression scores in the MCI group compared to controls on a hospital depression scale (Menon et al., 2021). These results suggest a contributory role of geriatric depression in impairing cognitive function in older adults (Mohan et al., 2019; Muhammad & Meher, 2021; Steffens & Potter, 2008).

### Factors influencing MoCA

Age, glycated hemoglobin levels, geriatric depression, and trait anxiety were factors that correlated negatively with MoCA performance. Of these, glycated hemoglobin and geriatric depression had a greater influence on MoCA performance than age or trait anxiety. Although it is known that metabolic dysregulation and geriatric depression can accelerate cognitive decline (Steffens and Potter, 2008; Sullivan et al., 2013; Yaffe, 2007), their contribution relative to age is an interesting finding from our study.

## 6. Limitations

This study has a relatively small sample to reflect community data. With the onset of the pandemic, additional data collection was not possible. However, we attempted to highlight the factors that have a with strong association with cognitive change in an elderly urban community. Since most cognitive tests were in English, and regional translations were unavailable for all tests, older adults with basic proficiency in English were included. However, most participants (>90%) were multilingual and therefore results from this study can be generalized to other multilingual, age and education matched, individuals.

## 7. Conclusions

Our study was an attempt to get an in-depth status of cognitive function in a general population of community dwelling older adults who had never visited a clinician for cognitive or mental health complaints. The participants were considered to be healthy and aging normally by themselves and their families, with no cognitive complaint. Using detailed cognitive assessments, we were able to profile MCI in approximately 17% (20/116) of our community dwelling older adult cohort. The relationship between cognitive function, age, glycated hemoglobin, and geriatric depression was highlighted. Performance in all cognitive domains declined as age advanced. High glycated hemoglobin levels were associated with lower processing speed and mild impairments in executive function. Vitamin B12 levels did not correlate to cognitive performance. Geriatric depression lowered performance on visuospatial tasks. Interestingly, glycated hemoglobin and geriatric depression levels were stronger predictors of changes in MoCA performance than age. Participants profiled as MCI also had higher levels of glycated hemoglobin. Taken together, our study has highlighted the significance of metabolic and psychological health for healthy cognitive aging. These observations are important for developing healthcare strategies in an increasingly diabetic nation like India.

## Data Availability

All data produced in the present work are contained in the manuscript

## 8. Acknowledgement

This study was supported by a fellowship from the Department of Science and Technology – Cognitive Science Research Initiative (DST-CSRI) awarded to BD. This work was also supported by a grant from the Pratiksha Trust awarded to AG, BD, and AS. Support from RIST (Rural India Support Trust) to AG and SH is also acknowledged.

## 9. Author Contributions

BD and AG conceptualized the study. BD and AS administered the tests on participants. SH coordinated serum data collection. BD and AS analyzed the data. All authors wrote the manuscript.

## 10. Conflict of Interest

The authors declare no conflict of interest.

## Notes

### Competing Interest Statement

The authors have declared no competing interest.

### Author Declarations

Institutional Ethics Committees at the National Center for Biological Sciences (NCBS) and the Institute of Ayurveda and Integrative Medicine Foundation for Revitalization of Local Health Traditions (IAIM FRLHT), both at Bengaluru gave ethical approval for this work.

